# Global, regional, and national burden of chronic kidney disease-related heart failure from 1990 to 2021: an analysis of data from the Global Burden of Disease Study 2021

**DOI:** 10.1101/2025.02.08.25321930

**Authors:** Jinzhong Ji, Feifan Chu, Qing Guan, Lumin Chen, Yuning Ma, Zujie Chen, Qiwei Ji, Mingxin Sun, Gaozhan Ren, Tingyang Huang, Haihan Song, Weiqiang Lin, Xiuquan Lin, Hao Zhou

**Author notes:** These authors contributed equally to this work (Jinzhong Ji, Feifan Chu, Qing Guan).

## Abstract

**Background:** The global burden of chronic kidney diseases related heart failure (CKD-related HF) has yet to be reported. The study aims to analyze the global burden of CKD-related HF.

**Methods:** Utilizing the data from the Global Burden of Disease Study 2021, we delineated the prevalence and years lived with disability (YLDs) of CKD-related HF at global, regional, and national levels, alongside its age and sex distributions, as well as temporal trends. Furthermore, we conducted transnational inequality analysis and frontier analysis. Additionally, we analyzed the burden of CKD-related HF with diverse etiological factors.

**Results:** Over the past three decades, the global burden of CKD-related HF has escalated, reaching 1,936,886 cases in 2021, with an age-standardized prevalence of 24.21 per 100,000 and an age-standardized YLDs rate of 3.07 per 100,000. This burden is projected to continue rising over the next decade. The burden of CKD-related HF is notably concentrated among the elderly and children. Notably, there is health inequity in its distribution, disproportionately affecting low sociodemographic index (SDI) groups, although alleviation opportunities exist across all SDI levels. Globally, the primary contributors to CKD-related HF are type 2 Diabetes Mellitus (T2DM) and hypertension, apart from CKD of other and unspecified causes.

**Conclusions:** The global burden of CKD-related HF has witnessed increase and is projected to persist over the next decade. Certain populations, including children, the elderly, and regions with low SDI levels, experienced a heavier burden. Effective management of primary diseases contributes to mitigating the burden of CKD-related HF.

---

**Keywords:** Chronic kidney disease, Heart failure, Prevalence, Years lived with disability, Global burden of disease

## Introduction

Chronic kidney diseases (CKD) are witnessing a gradual rise in incidence and mortality rates globally, imposing substantial medical and economic burdens on societies and healthcare systems.^1^ Cardiovascular disease stands as the primary risk factor and the leading cause of mortality among CKD patients, with heart failure (HF) being one of the most prevalent cardiovascular outcomes in these individuals.^2^

According to reports, the incidence of newly diagnosed HF in the general global population does not exceed 1%, whereas among CKD patients, it ranges from 17% to 21%.^3^ Data from the United States Renal Data System in 2020 revealed a prevalence of HF in CKD patients at 25.1%, compared to 6.6% in non-CKD individuals. Notably, the prevalence of HF increases with advancing CKD stages: 21.5% in stages 1-2, 28.4% in stage 3, and 41.3% in stages 4-5.^4^ CKD patients with coexisting HF exhibit poorer quality of life and heightened mortality risks compared to those with either condition alone.^5^ CKD-related HF patients represent a complex syndrome, underscoring the substantial and urgent unmet medical needs in this population. However, prior research has predominantly been confined to individual countries or regions.^6^ A comprehensive study on the global burden of CKD-related HF is yet to emerge, and there remains a paucity of comprehensive epidemiological data on this issue.

The Global Burden of Diseases, Injuries, and Risk Factors Study (GBD) 2021 is a comprehensive study of global health loss.^7^ In this study, we conducted a systematic analysis of the global burden of CKD-related HF utilizing data from the GBD 2021. This analysis aims to provide evidence to guide public health strategies and resource allocation to reduce the burden of CKD-related HF.

## Methods

### Case definition and data sources

According to the GBD 2021, CKD is denoted by a permanent loss of kidney function, as indicated by the estimated glomerular filtration rate and the urine albumin-to-creatinine ratio. The corresponding ICD-10 for CKD include N18.1 through N18.9.^7^

HF is described as a complex clinical syndrome resulting from structural or functional abnormalities of the heart, which lead to impaired ventricular contraction or filling.^8^ Within the GBD 2021 data sources, HF is classified as a health impairment and is diagnosed using structured, symptom-based criteria such as the Framingham or European Society of Cardiology standards. The criteria established by the American College of Cardiology and the American Heart Association for stage C and above are utilized to identify both currently symptomatic patients and those diagnosed with HF who are asymptomatic at present.^7^

In the context of the GBD 2021, CKD-related HF refers specifically to heart failure attributed to volume overload resulting from CKD.^9^ For GBD 2021,only CKD Stage 5 is attributed a disability weight associated with heart failure. The classification of CKD as a causative factor for HF is established through literature reviews and expert consensus, based upon the overall prevalence of HF and subsequently estimating the prevalence of CKD-related HF according to the proportion attributable to CKD.^7^ Detailed methodologies for these calculations have been previously published in the Lancet.^7^

In this study, we obtained estimates of the number of cases, prevalence, YLDs, and YLDs rate of CKD-related HF, along with their 95% UIs, from the GBD 2021 website. This study adheres to the Guidelines for Accurate and Transparent Health Estimates Reporting statement.

### Data Adjustment and Statistical Analysis

Firstly, an analysis of the age-standardized prevalence (ASPR) and YLDs rate of CKD-related HF was conducted to delineate its distribution across various global regions and countries. To further dissect the age and sex disparities in CKD-related HF, the disease burden was quantified by stratifying the data by both sex and age. Additionally, a Spearman correlation analysis was employed to examine the relationship between the Sociodemographic Index (SDI) and the age-standardized ASPR and YLDs rate, thereby elucidating the association between the burden of CKD-related HF and SDI development.

The temporal trends of CKD-related HF globally and across varying SDI regions from 1990 to 2021 were analyzed utilizing the Joinpoint regression model. This model meticulously computes the Annual Percent Change (APC) alongside its corresponding 95%UI.^10^ Furthermore, we have utilized the Bayesian Age-Period-Cohort (BAPC) model to delineate and forecast the trends of heart failure among CKD patients up to the year 2031^11^.

In the GBD 2021, the classifications of CKD include CKD attributed to Type 1 Diabetes Mellitus (T1DM), CKD attributed to Type 2 Diabetes Mellitus (T2DM), CKD resulting from glomerulonephritis, CKD caused by hypertension, and CKD of other and unspecified causes (including hereditary nephropathy, cystic kidney disease, congenital obstructive defects of renal pelvis, and congenital malformations of the ureter, among others). We conducted an analysis of the distribution of CKD-related HF burden attributable to different types of CKD across regions, age groups, sexs, and time periods.

In this study all statistical analyses and data visualizations were conducted using the R software package.

## Result

### CKD-related HF Burden: A Geographic Analysis

Globally, in 2021, the number of cases of CKD-related HF amounted to 1,936,886 (95% UI: 1,600,208 to 2,343,466). The ASPR was 24.21 cases per 100,000 (95% UI: 19.86 to 29.23). From 1990 to 2021, the percentage change in ASPR was 78.23%(95% UI, 69.87% to 86.79%) (Table S1). Additionally, the YLDs due to CKD-related HF totaled 245,658 globally (95% UI: 154,573 to 356,658), with an age-standardized YLDs rate of 3.07 per 100,000 (95% UI: 1.95 to 4.44). Over the period from 1990 to 2021, the percentage change in the YLDs rate was 77.61 (95% UI, 69.31% to 86.28%) (Table S1).

Regionally, Western Sub-Saharan Africa exhibited the highest ASPR for CKD-related HF in 2021, with 90.21 cases per 100,000 (95% UI: 69.91 to 113.99), followed by Andean Latin America at 62.49 cases per 100,000 (95% UI: 50.13 to 75.79) (Table S1). In contrast, Eastern Europe had the lowest ASPR, with 6.04 cases per 100,000 (95% UI: 4.50 to 7.80), followed by Oceania, with 10.97 (95% UI: 8.75 to 14.16) cases per 100,000. From 1990 to 2021, the ASPR varied across regions, but an overall upward trend was observed. Notably, High-income North America demonstrated the most substantial increase, with an ASPR rise of 1.99 cases per 100,000 (95% UI: 1.56 to 2.49). Similar to ASPR, the age-standardized YLDs rate for CKD-related HF also showed an upward trend across regions. High-income North America again led in terms of the magnitude of increase(Table S1).

At the national level, Nigeria, El Salvador, and Nicaragua emerged as the top three countries with the highest ASPR and age-standardized YLDs rate for CKD-related HF in 2021, while Ukraine, Belarus, and Iceland recorded the lowest rates (Figure 1, Table S2). Additionally, we analyzed the burden of CKD-related HF in five representative countries categorized by their SDI levels: America, Republic of Turkey, China, India, and Pakistan, listed from highest to lowest SDI. Among these countries, China has the highest number of CKD-related HF cases, followed by India and America. However, in terms of prevalence, Republic of Turkey exhibits the highest rate, followed closely by America; both countries have prevalence rates that exceed the global average. A similar trend is observed in YLDs. Over the period from 1990 to 2021, the ASPR in these countries has been increasing, with Republic of Turkey experiencing the most rise, followed by America and Pakistan, and YLDs reflecting the same trend (Figure S1, Table S2).

**Figure 1.**
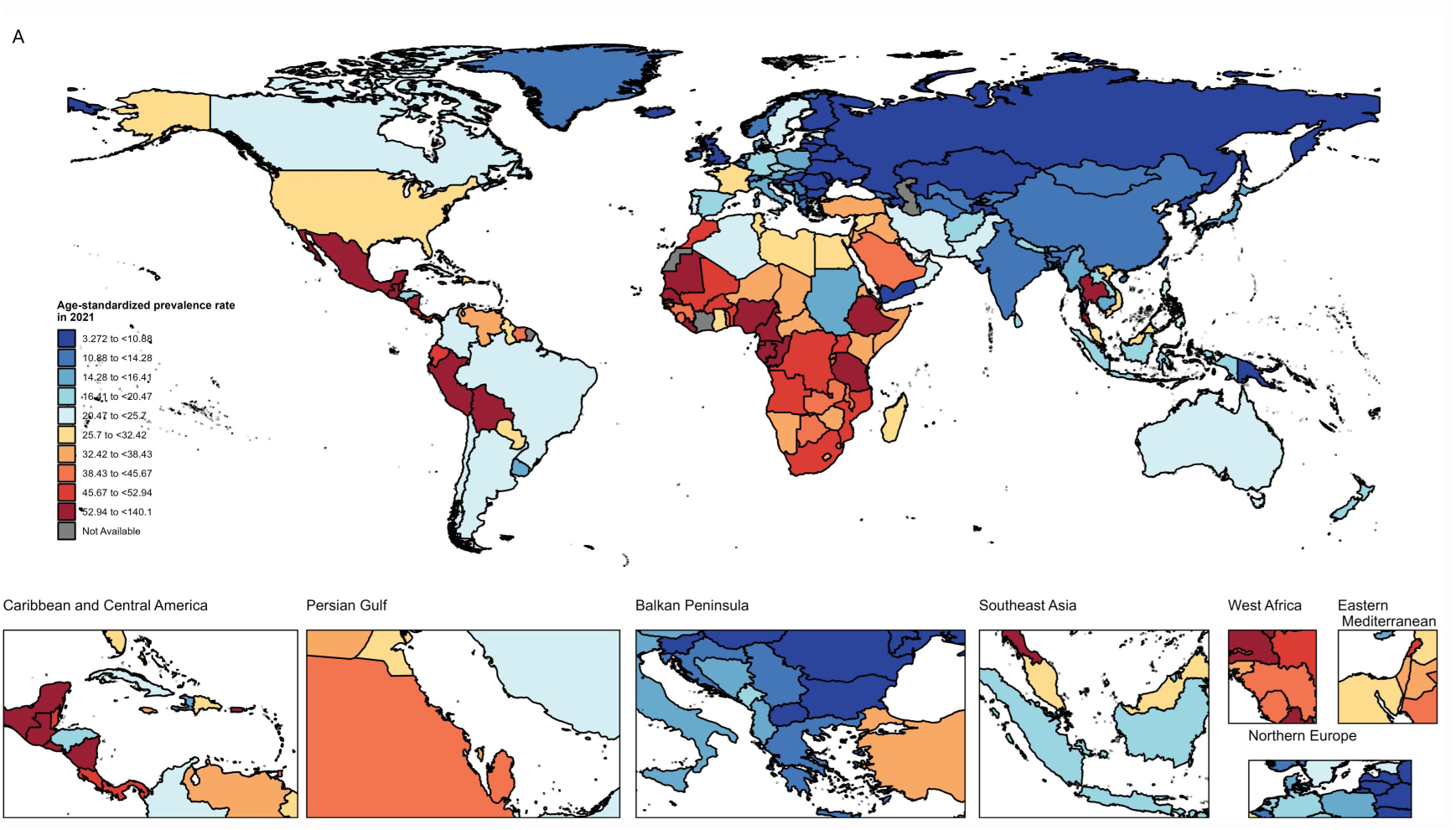
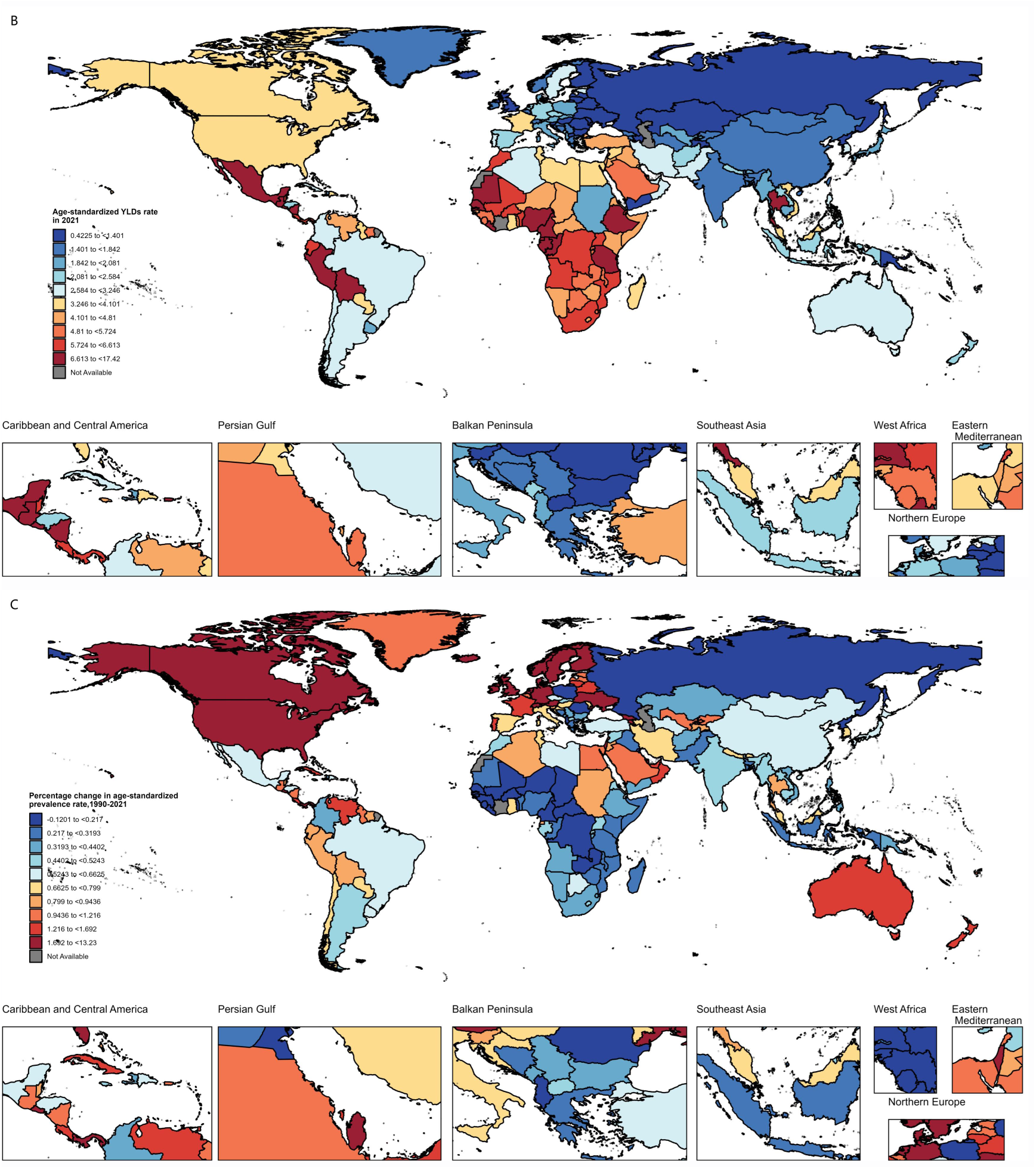
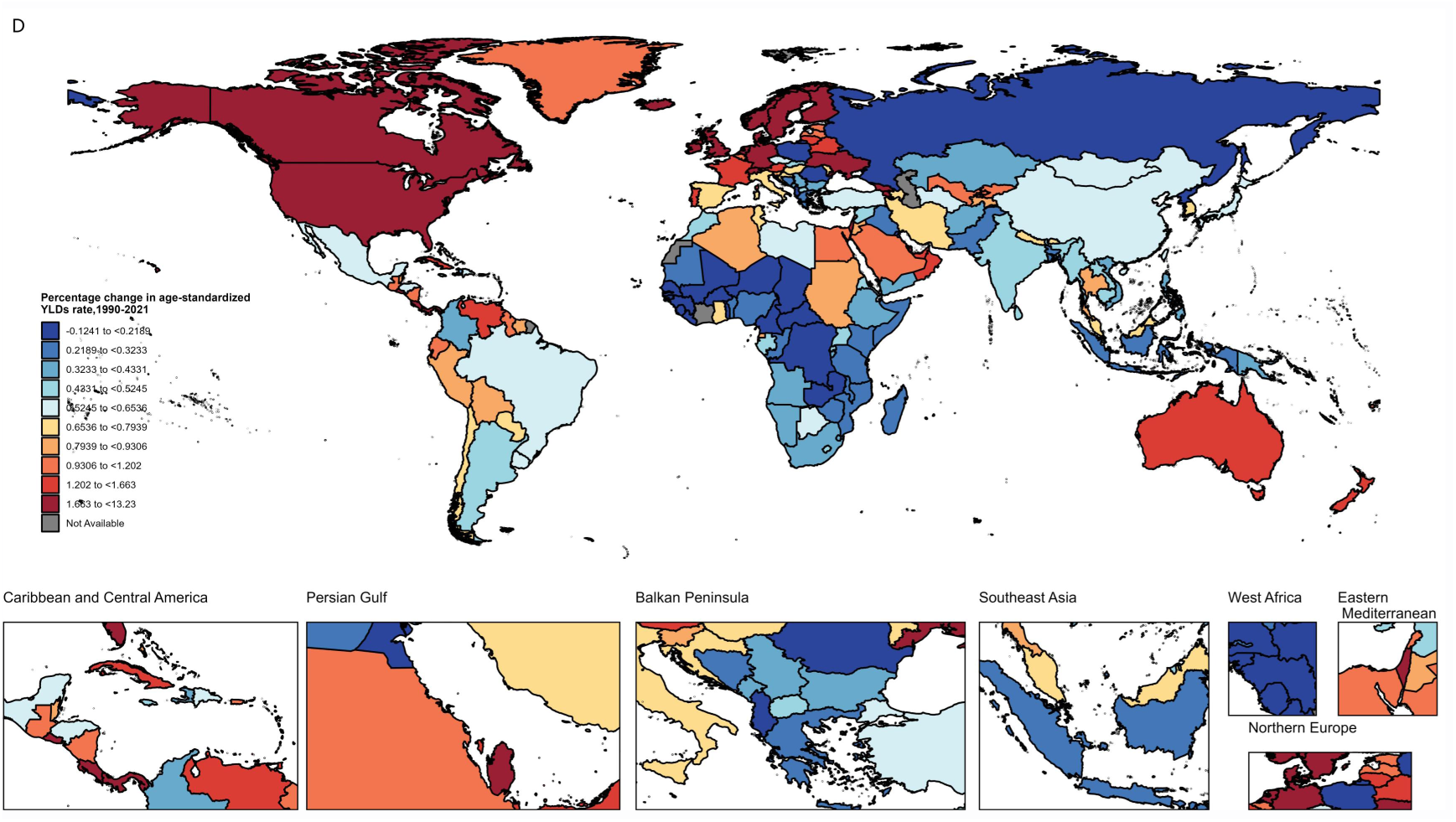
Geographical distribution of the burden of CKD-related HF. ASPR (A)and Age-standardized YLDs rate (B) per 100,000 in 2021; Percentage change in ASPR (C) and age-standardized YLDs rate (D) per 100,000 from 1990 to 2021.

### Age and sex Patterns of CKD-related HF Burden

In 2021, the highest number of prevalent cases of CKD-related HF globally was observed in the 70-74 age group, with 239,141.8 cases (95% UI: 145,682.4 to 381,046.9), followed closely by the 65-69 age group, accounting for 198,308.5 cases (95% UI: 121,333 to 309,648.3). Notably, substantial case numbers were also reported in the under 5 age group (151,853.5 cases, 95% UI: 101,039.3 to 222,179.9) and the 5-9 age group (138,721.8 cases, 95% UI: 77,323.6 to 233,352.3) (Table S3). Regarding prevalence rates, for both sex within each 5-year age group, individuals under the age of 30 exhibit a decrease in prevalence as age advances. Conversely, for those over the age of 30, prevalence tends to rise with increasing age (Figure 2A, Table S3). A similar trend was observed in YLDs (Figure 2B, Table S4). The aforementioned outcomes indicate that the burden of CKD-related HF is notably concentrated among the elderly and children.

**Figure 2.**
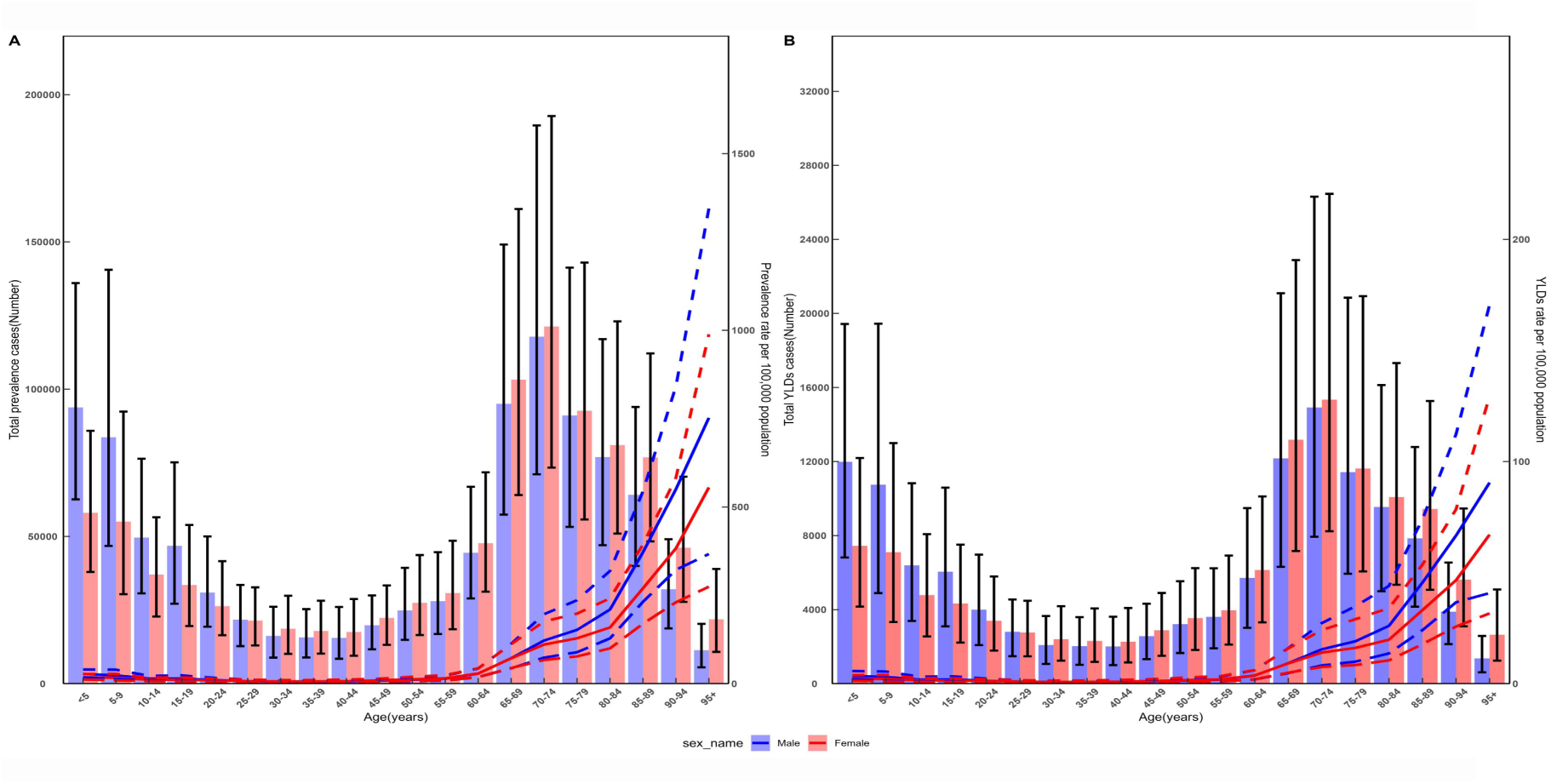
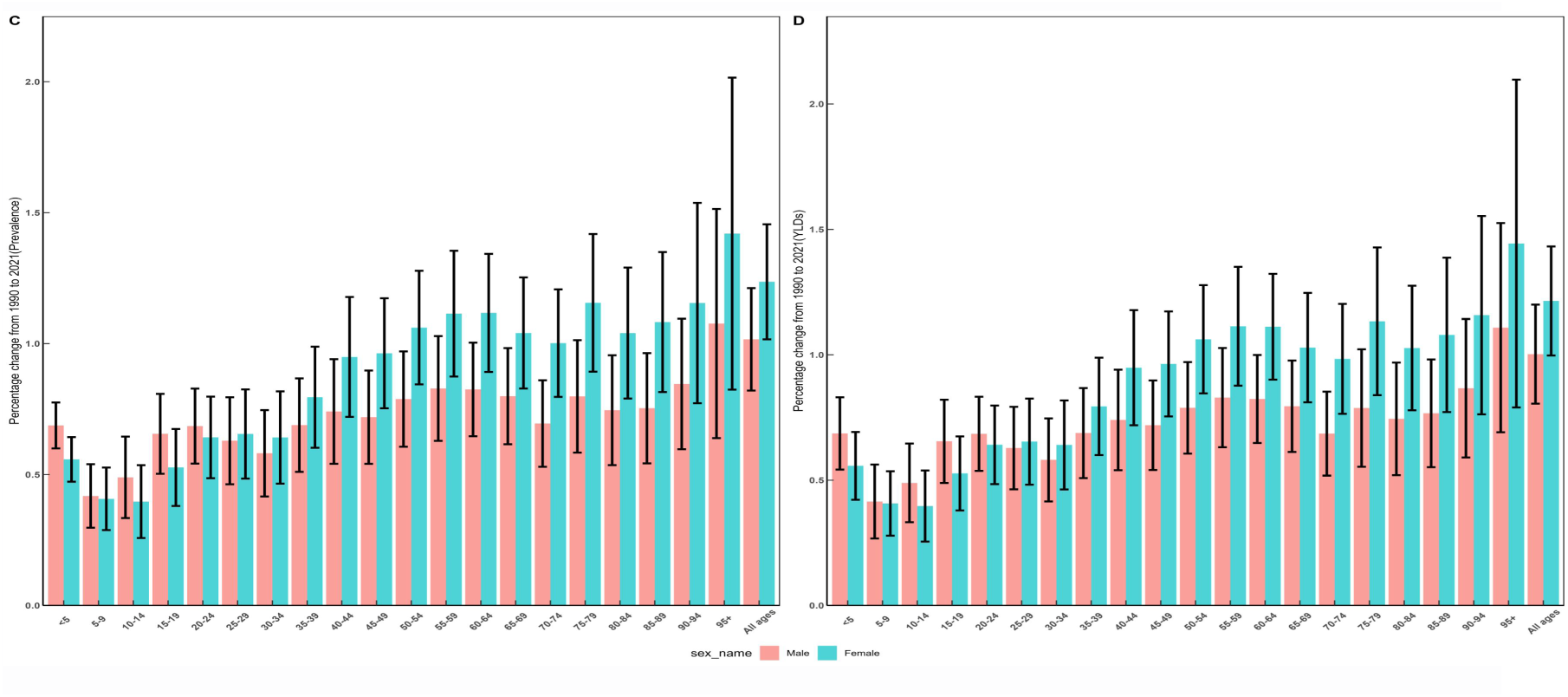
Age and sex patterns of CKD-related HF burden. (A) Global CKD-related HF prevalent cases and ASPR per 100,000 in 2021; (B) Global CKD-related HF YLDs and Age-standardized YLDs rate per 100,000 in 2021; Percentage change in ASPR (C) and Age-standardized YLDs rate (D) per 100,000 from 1990 to 2021.

Remarkably, regarding sex distribution, females surpassed males in case counts above 30 years of age, whereas males predominated in the under-30 age bracket. This sex-specific pattern was mirrored in the distribution of YLDs (Figure 2A, Table S3-4). From a prevalence perspective, no significant sex disparity was evident below 65 years, whereas males exhibited a higher prevalence than females above 65 years. A similar trend was observed in the YLDs rate (Figure 2A-B, Table S3-4).

Between 1990 and 2021, the age-standardized prevalence and YLD rate (per 100,000 individuals) of CKD-related HF increased across all age groups, with the most pronounced growth observed in the 95+ age group. Intriguingly, when analyzed by sex, females aged 25 and above demonstrated a greater increase in both age-standardized prevalence and YLD rate compared to males (Figure 2C-D, Table S3-4).

### Temporal Trends in the Burden of CKD-related HF

Joinpoint regression analysis reveals an overall upward trend in the ASPR of CKD-related HF globally from 1990 to 2021, with an Annualized Average Percentage Change (AAPC) of 1.89%. Notably, the most pronounced increase occurred between 1995 and 2003 (APC = 2.75%), while the smallest rise was observed from 2018 to 2021 (APC = 0.48%) (Table S5). A similar pattern was discerned in age-standardized YLDs (Table S5).

When stratified by SDI regions, the ASPR and YLDs rate of CKD-related HF also demonstrated an overall upward trend from 1990 to 2021. Notably, regions with higher SDI exhibited more pronounced changes. Specifically, the High SDI regions exhibited the most substantial increase in both ASPR (AAPC = 2.86%) and age-standardized YLDs (AAPC = 2.82%). Conversely, Low SDI regions exhibited the smallest changes in ASPR and age-standardized YLDs (AAPC = 0.72% for both) (Figure S2, Table S5).

The results of the Bayesian Age-Period-Cohort (BAPC) model analysis project that the ASPR of CKD-related HF will increase from 24.21 per 100,000 in 2021 to 28.31 per 100,000 by 2031. Similarly, the predicted age-standardized YLDs rate is anticipated to continue rising over the next decade, from 3.07 per 100,000 in 2021 to 3.46 per 100,000 by 2031 (Figure 3A-B, Table S6).

**Figure 3.**
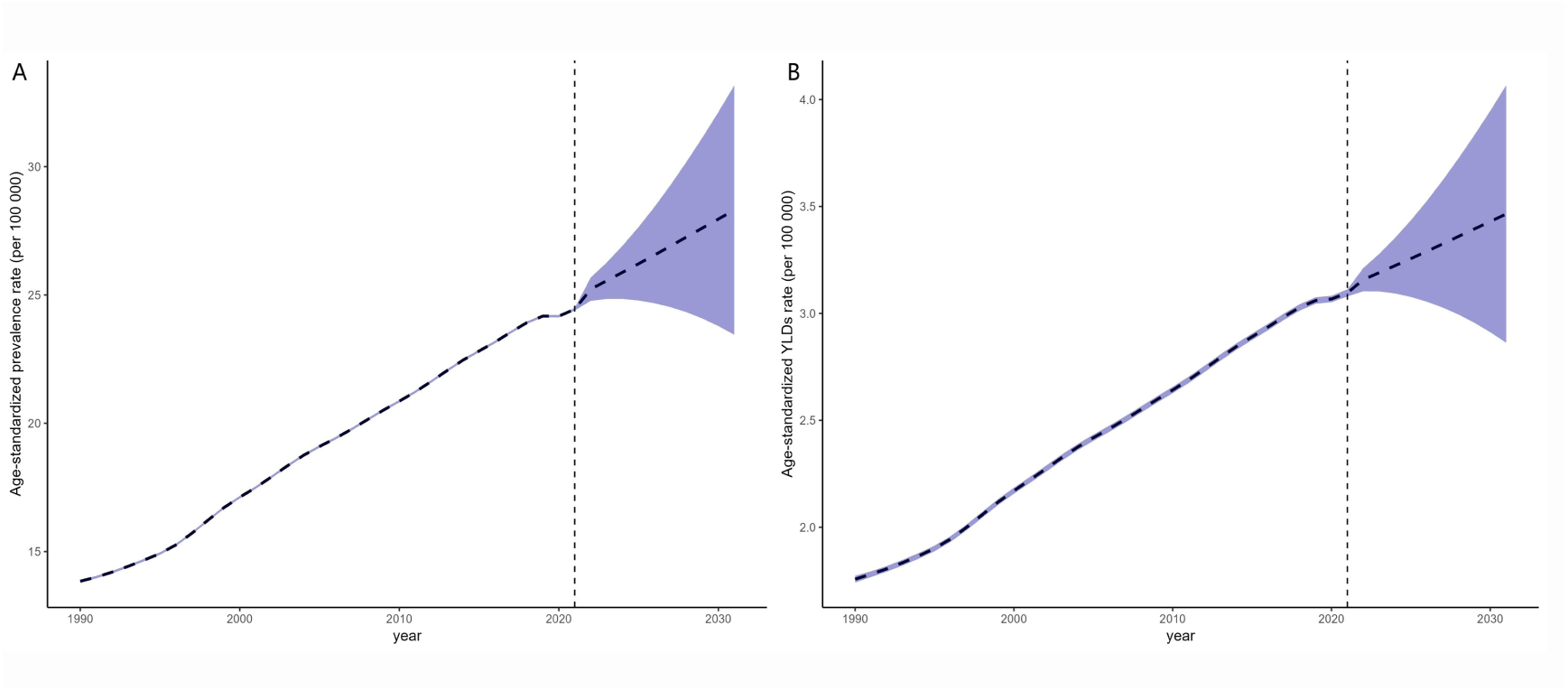
Trends in the burden of CKD-related HF over the next 10 years. Prediction of global ASPR (A) and age-standardized YLDs Rate (B) per 100,000 of global CKD-related HF.

### CKD-related HF Burden Stratified by SDI

When analyzing the CKD-related HF burden across various SDI regions, it is evident that Low SDI regions exhibit the highest ASPR, with 38.71 cases per 100,000 individuals (95% UI: 30.16 to 49.45). In contrast, regions with higher income levels, including High-middle SDI and High SDI, demonstrate lower ASPR of 15.08 (95% UI: 12.60 to 18.41) and 22.16 (95% UI: 17.83 to 27.14) cases per 100,000, respectively (Table S1). A similar pattern is observed in the age-standardized YLDs rate.

Overall, a negative correlation is evident between SDI and the burden of CKD-related HF, with a correlation coefficient of -0.378 (P<0.0001) for ASPR. Notably, the observed global ASPR for CKD-related HF are slightly lower than anticipated. Regionally, from 1990 to 2021, the observed ASPR in Western Sub-Saharan Africa, Andean Latin America, Southern Sub-Saharan Africa, and Central Latin America surpassed their SDI-based expectations (Figure 4A). At the national level, based on SDI, certain countries such as Nigeria, El Salvador, and Nicaragua had significantly higher observed ASPR than predicted, whereas Tajikistan, Papua New Guinea, and Afghanistan had rates far below expectations (Figure 4B). A similar pattern is observed in the age-standardized YLDs rate (Figure S3).

**Figure 4.**
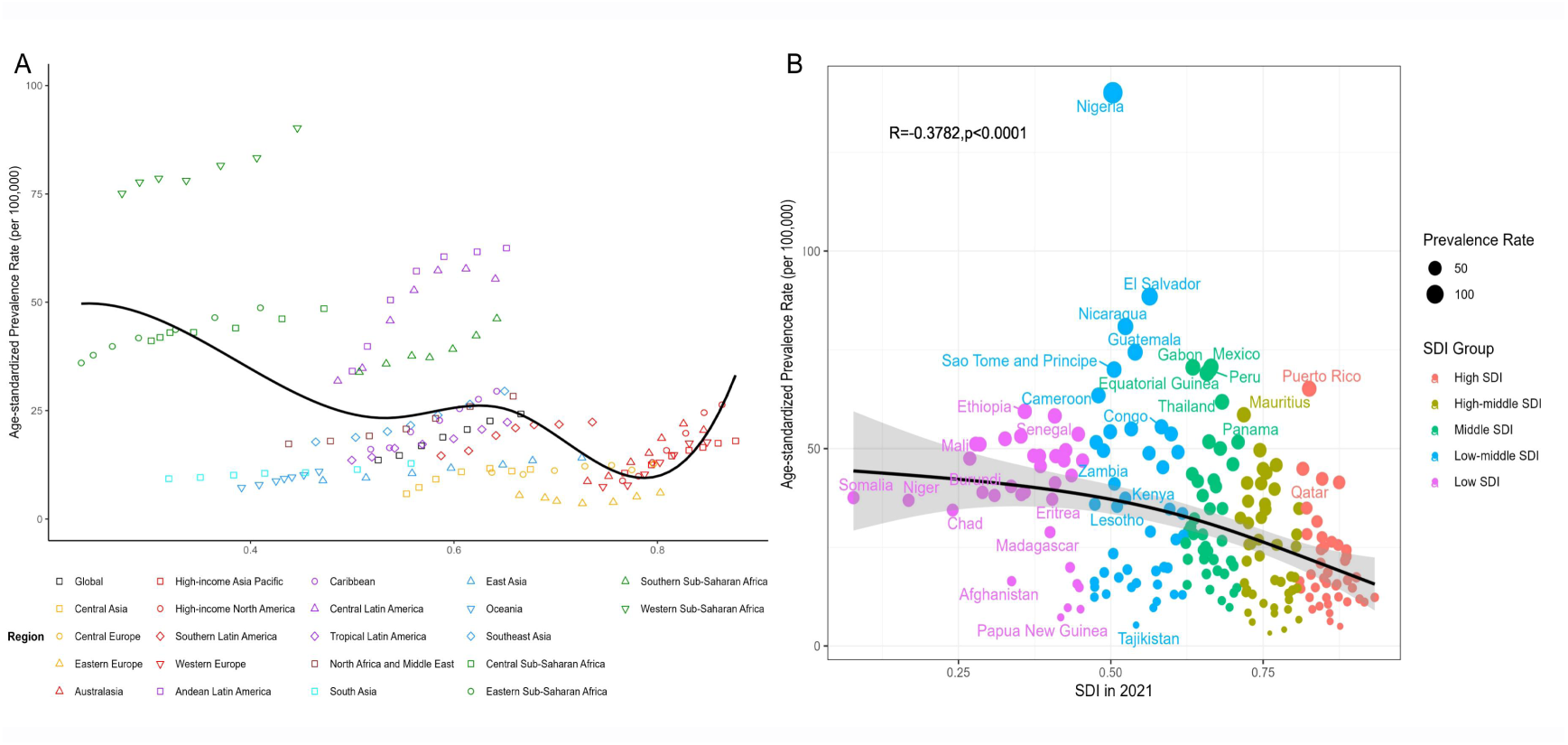
Association between CKD-related HF burden and SDI. Association between age-standardized prevalence of CKD-related HF and SDI by 21 GBD regional (A), and by the national level (B).

Within the context of the burden posed by CKD-related HF, absolute and relative inequalities associated with SDI have been observed, with nations at the lower end of the SDI spectrum shouldering a disproportionately high load. The SII reveals a narrowing gap in the ASPR of CKD-related HF between countries with the highest and lowest SDIs, decreasing from -26.33 in 1990 to -20.80 in 2021 (Figure 5A, Table S7), indicating a reduction in health inequalities. The concentration curves demonstrate that, in both 1990 and 2021, the majority of the ASPR for CKD-related HF lies above the line of equality, signifying a concentration of the ASPR’s geographical distribution in impoverished regions (Figure 5B, Table S7).

**Figure 5.**
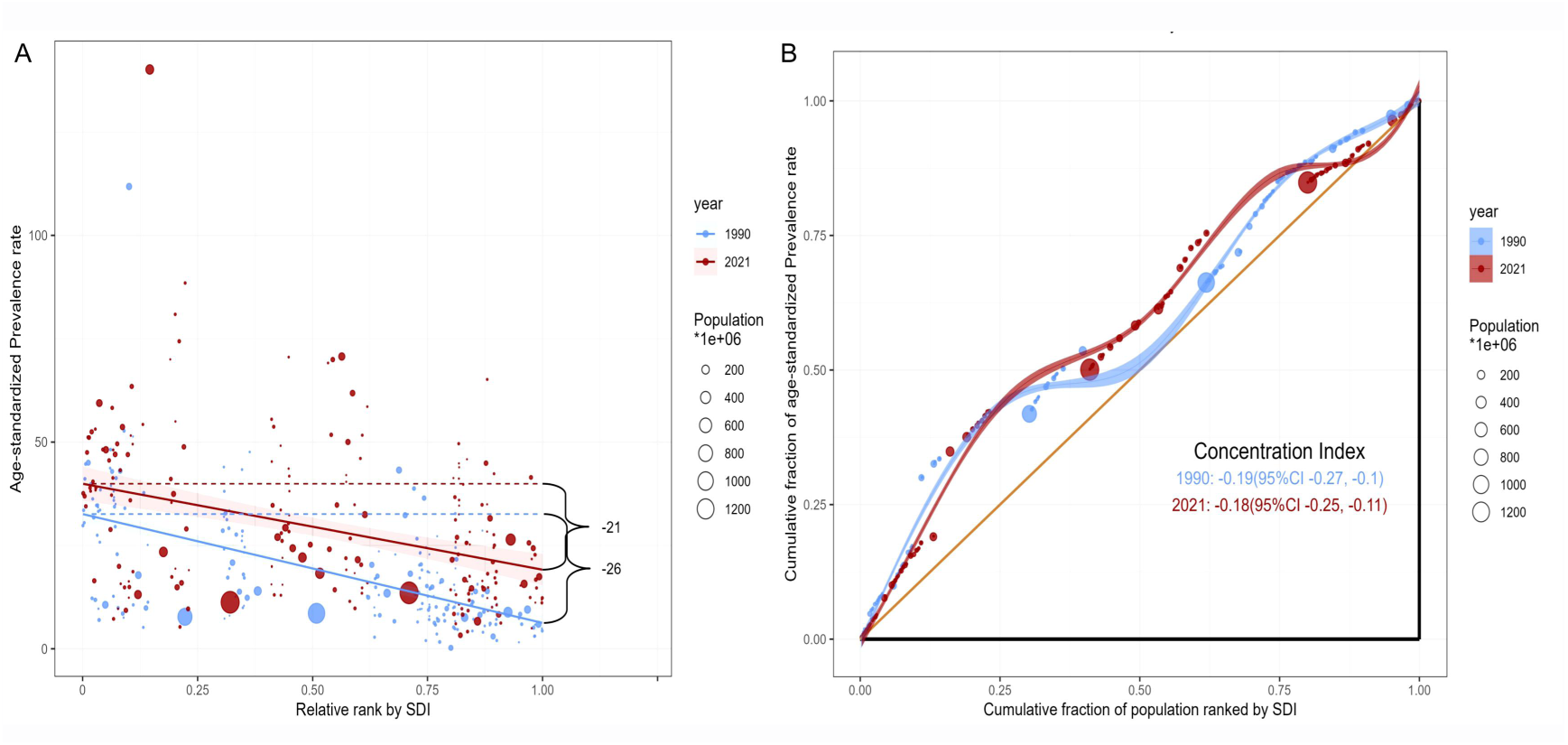
Cross-country inequality analysis of CKD-related HF burden. Health inequality regression curve (A) and concentration curves (B) for age-standardized prevalence rates of CKD-related HF in 1990 and 2021.

Furthermore, the gap in the age-standardized YLDs rate due to CKD-related HF between the highest and lowest SDI countries has narrowed from -3.27 in 1990 to -2.58 in 2021 (Figure S4A, Table S7). Similarly, the concentration curves for the age-standardized YLDs rate in 1990 and 2021 both reside above the line of equality (Figure S4B, Table S7).

### Frontier Analysisof CKD-related HF Burden Based on Socio-Demographic Indices

Overall, the effective disparity observed in both the ASPR due to CKD-related HF has tended to widen within the given SDI range across the period from 1990 to 2021, with an increasing burden in most countries (Figure 6, Table S8). Minimal effective disparity is noted in countries with higher SDI; however, the highest effective disparity is observed in those situated within the middle SDI spectrum (Figure 6A, Figure S5A).

**Figure 6.**
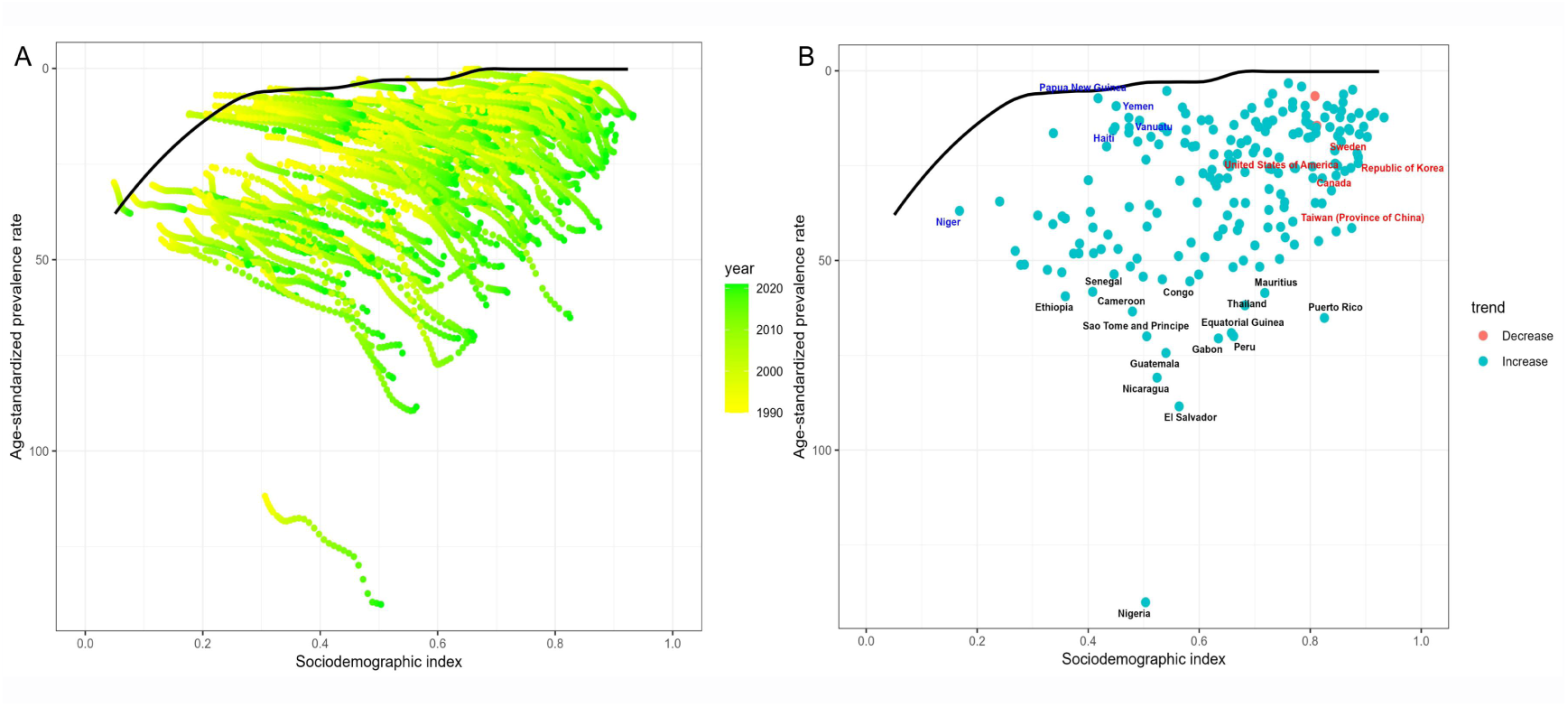
Frontier analysis based on the SDI and the burden of CKD-related HF. Frontier analysis of the ASPR of CKD-related HF and SDI from 1990 to 2021 (A), and in 2021 (B). Note: The top 15 countries with the greatest effective disparities are marked in black. Regions with high Socio-demographic Index (SDI) values (>0.85) and substantial effective disparities are highlighted in red, while those with low SDI values (<0.5) and smaller effective disparities are indicated in blue. The boundary, depicted in pure black, represents the potential age-standardized prevalence achievable based on SDI. Actual values for countries and regions are denoted by dots, with increases in the burden of heart failure due to central CKD from 1990 to 2021 represented by blue dots and decreases by red dots.

Regarding the ASPR, Nigeria, El Salvador, and Nicaragua emerge as the three countries most likely to exhibit a narrowing of the disparity. Among nations with lower SDI, Papua New Guinea and Yemen exhibit minimal effective disparity, whereas countries with higher SDI, such as Sweden, and the United States of America, demonstrate larger disparities (Figure 6B, Table S8).

For the age-standardized YLDs rate, Nicaragua, Guatemala, and Mexico are the three countries poised to potentially narrow the disparity gap. Notably, some low SDI countries yield favorable outcomes, with Somalia and Papua New Guinea, both with lower SDI, displaying minimal effective disparity. Conversely, disappointment arises from the larger disparities observed in countries with higher SDI, including Canada, the United States of America, and the Republic of Korea (Figure S5B, Table S8).

### Distribution of CKD-related HF of Diverse Etiologies

In 2021, the primary etiological contributors to CKD-related HF population were CKD of other and unspecified causes, T2DM, and hypertension in global (Figure S6A-B). This global trend is mirrored across varying SDI regions, CKD of other and unspecified causes and T2DM consistently emerging as the leading etiological factors.

Stratifying by age groups, CKD of other and unspecified causes predominates as the primary etiology underlying the CKD-related HF burden across all age cohorts. Among individuals aged over 85, regardless of sex, hypertension induced CKD ranks as the second leading cause of CKD-related HF burden. In contrast, for age groups ranging from 0-5 years and 45-85 years (excluding the 50-54 years age bracket), T2DM induced CKD emerges as the second most prevalent etiology. Notably, within the 5-44 years age group, glomerulonephritis assumes the position of the second most common cause of CKD (Figure S6C-F). From 1990 to 2021, CKD attributed to Other Causes, T2DM, and hypertension have collectively served as the primary drivers of the escalating HF burden within the global CKD population (Figure S6G-H).

## Discussion

In 2021, the ASPR of CKD-related HF was highest in Western Sub-Saharan Africa, whereas it was lowest in Eastern Europe. Intriguingly, despite the reported highest prevalence of CKD globally in Eastern and Central Europe,^15^ the ASPR of CKD-related HF patients in these regions was the lowest, potentially attributable to their adequate access to healthcare services.^16^ Notably, High-income North America exhibited the most substantial upward trend. At the national level, the top three countries with the highest ASPR in 2021 were Nigeria, El Salvador, and Nicaragua. This may stem from a lack of prioritization for CKD-related HF and inadequate medical resources in these nations.^17^

The burden of CKD-related HF is disproportionately distributed across different age groups, with over half of the cases occurring in individuals aged over 60 years. Consistent with previous studies, both the age-standardized prevalence and YLDs rate of CKD-related HF significantly increase with advancing age, particularly among those aged 65 and above. The global increase in the number of CKD-related HF cases and prevalence can be attributed to the aging of the population^18^. It is noteworthy that significant numbers of cases and YLDs are also observed in children. This discrepancy may be attributed to the ongoing development of the cardiovascular system in children, which results in a relatively weaker capacity of the heart to adapt to stress and volume overload. Consequently, children are more susceptible to the effects of fluid overload and electrolyte imbalances induced by chronic kidney disease,^19^ leading to an increased incidence of HF. However, additional investigations are warranted to explore the more nuanced underlying factors contributing to this phenomenon. In conclusion, the burden of CKD-related HF warrants attention not only among the elderly but also in children.

Our study found that males exhibited a higher age-standardized prevalence and YLDs than females in above 65 years. These sex differences in CKD-related HF may be partly attributed to conventional risk factors such as aging, hypertension, and smoking.^20^ Moreover, encompassing sex identity, health care access and income inequalities, may also contribute to the observed sex disparities in CKD-related HF burden.^21^ Consequently, there is a pressing need to develop sex-specific management strategies to more precisely address CKD-related HF.

The burden of heart failure within the context of CKD exhibits inequities across different SDI regions. The cross-national inequality analysis identified that the burden of CKD-related HF is predominantly concentrated in impoverished areas. Analogously, in the United Kingdom, socioeconomically disadvantaged populations are more prone to heart failure compared to their wealthier counterparts.^22^ The preponderance of the burden of chronic kidney disease is centered in low-middle and low SDI regions, which may underlie the heavier burden of CKD-related HF observed in Low SDI regions.^23^ Additionally, poor adherence to cardiovascular medications, unavailability, or unaffordability in low and middle SDI countries represent potential contributing factors.^24^ Notably, the disparity in the burden of CKD-related HF across SDI regions is diminishing, potentially attributable to the notably greater increase in age-standardized prevalence and YLDs rate in High SDI regions than in low SDI regions from 1990 to 2021. The rapid growth in the burden in High SDI regions may stem from more pronounced population growth and aging levels.^25^ Frontier analysis demonstrates considerable heterogeneity in effective differences among SDI countries, with the most pronounced differences observed in middle-range SDI nations, yet opportunities exist across all levels of the SDI to mitigate this burden. In conclusion, given the limited medical resources in underdeveloped countries, governments in these nations should endeavor to mitigate the burden of this disease through enhanced early screening, and health literacy promotion.

Our findings indicate that, excluding CKD of other and unspecified causes, CKD arising from T2DM and hypertension constitutes the primary source of the global burden of CKD-related HF. Both diabetes and hypertension are common risk factors for both CKD and HF. Hyperglycemia can lead to microvascular pathology, thereby facilitating the progression of heart failure,^25^ while also elevating levels of reactive oxygen species, advanced glycation end-products and so on, causing vasoconstriction and exacerbating CKD.^26^ Hypertension is a pivotal determinant of adverse renal and cardiovascular outcomes. Renal dysfunction elevates blood pressure through sodium retention and activation of the RAAS and the sympathetic nervous system, which in turn exacerbates CKD, perpetuating a vicious cycle.^27^ Additionally, Sustained hypertension leads to progressive diastolic dysfunction, decreased left ventricular filling due to concentric remodeling, and ultimately, the development of heart failure.^28^ Consequently, controlling the burden of CKD-related HF also necessitates a focused effort on the prevention and treatment of primary conditions.

This study has several limitations. Firstly, it is subject to the common constraints inherent in all GBD studies, particularly with respect to the heterogeneity of data collection methods and sources, as well as the quality and completeness of data. Secondly, Data collection on CKD-related HF in economically disadvantaged regions is often limited due to inadequate reporting and diagnosis. Consequently, the GBD project uses statistical methods to estimate CKD-related HF burdens in data-scarce regions, providing estimates with appropriate uncertainty rather than omitting critical information. Finally, since the GBD 2021 study did not consider heart failure as a potential cause of death, we were unable to assess deaths and disability-adjusted life years attributed to CKD-related HF.

In summary, the burden of CKD-related HF have increased globally and are projected to continue rising over the next decade. This burden is notably concentrated among the elderly and children. Regional inequalities in the burden of CKD-related HF persist, particularly in low SDI levels regions and countries, yet opportunities exist across all levels of the SDI to mitigate this burden. Preventing and managing primary conditions such as type 2 diabetes and hypertension are crucial in controlling CKD-related HF. Consequently, policymakers should judiciously allocate public health resources to mitigate the challenging burden of CKD-related HF.

## Contributors

HZ and XQL conceived and designed the study. JZJ, FFC and LMC drafted the manuscript. QG, YNM, QWJ, MXS, and GZR analyzed the GBD data. HZ, HHS, TYH, and XQL were responsible for the article checking and reviewing. HHS, XQL, and JJZ accessed and verified the data. All authors revised the report and approved the final version before submission. HZ had access to all the data in the study and had final responsibility for the decision to submit for publication.

## Data Availability

All data produced in the present study are available upon reasonable request to the authors

https://ghdx.healthdata.org/gbd-2021/results

## Data sharing

The data of this study are publicly available at the GBD 2021 website (https://ghdx.healthdata.org/gbd-2021/results). All data also will be made available on request to the corresponding author.

## Acknowledgments

We highly appreciate the work by the GBD 2021 collaborators.

## References

1 Francis A, Harhay MN, Ong ACM, et al. Chronic kidney disease and the global public health agenda: an international consensus. Nat Rev Nephrol. 2024;20(7):473–485.

2 House AA, Wanner C, Sarnak MJ, et al. Heart failure in chronic kidney disease: conclusions from a Kidney Disease: Improving Global Outcomes (KDIGO) Controversies Conference. Kidney Int. 2019;95(6):1304–1317.

3 Romero-González G, Ravassa S, González O, et al. Burden and challenges of heart failure in patients with chronic kidney disease. A call to action. Nefrologia (Engl Ed). 2020;4 0(3):223–236.

4. https://usrds-adr.niddk.nih.gov/2020/chronic-kidney-disease/4-cardiovascular-disease-in-patients-withckd

5 Saran R, Robinson B, Abbott KC, et al. US Renal Data System 2016 Annual Data Rep ort: Epidemiology of Kidney Disease in the United States. Am J Kidney Dis. 2017;69(3 Suppl 1):A7–A8.

6 Shearer JJ, Hashemian M, Nelson RG, et al. Demographic trends of cardiorenal and heart failure deaths in the United States, 2011-2020. PLoS One. 2024;19(5):e0302203.

7 GBD 2021 Diseases and Injuries Collaborators. Global incidence, prevalence, years lived with disability (YLDs), disability-adjusted life-years (DALYs), and healthy life expectancy (HALE) for 371 diseases and injuries in 204 countries and territories and 811 subnational locations, 1990-2021: a systematic analysis for the Global Burden of Disease Study 202 1. Lancet. 2024;403(10440):2133–2161.

8 Heidenreich PA, Bozkurt B, Aguilar D, et al. 2022 AHA/ACC/HFSA Guideline for the Management of Heart Failure: A Report of the American College of Cardiology/American Heart Association Joint Committee on Clinical Practice Guidelines [published correction appears in J Am Coll Cardiol. 2023 Apr 18;81(15):1551. doi: 10.1016/j.jacc.2023.03.002]. J A m Coll Cardiol. 2022;79(17):e263-e421.

9. Institute for Health Metrics and Evaluation (IHME). GBD 2021 Cause and Risk Summary: [Heart failure]. Accessed [2024/5/31]). Seattle, USA: IHME, University of Washington, 2024.

10 Tuo Y, Li Y, Li Y, et al. Global, regional, and national burden of thalassemia, 1990-20 21: a systematic analysis for the global burden of disease study 2021. EClinicalMedicine. 2024;72:102619.

11 Li C, Hua G, Liu S, Yu H, Yang X, Liu L. Global, regional, and national burden of blindness and vision loss attributable to high fasting plasma glucose from 1990 to 2019, and forecasts to 2030: A systematic analysis for the Global Burden of Disease Study 201 9. Diabetes Metab Res Rev. 2024;40(4):e3802.

12 Organization WH. Handbook on health inequality monitoring with a special focus on low and middle-income countries: Handbook on health inequality monitoring with a special focus on low and middle-income countries.;2013.

13 Lu M, Li D, Hu Y, et al. Persistence of severe global inequalities in the burden of Hypertension Heart Disease from 1990 to 2019: findings from the global burden of disease study 2019. BMC Public Health. 2024;24(1):110. Published 2024 Jan 6.

14 Bai Z, Han J, An J, et al. The global, regional, and national patterns of change in the burden of congenital birth defects, 1990-2021: an analysis of the global burden of diseas e study 2021 and forecast to 2040. EClinicalMedicine. 2024;77:102873.

15 Bello AK, Okpechi IG, Levin A, et al. An update on the global disparities in kidney disease burden and care across world countries and regions. Lancet Glob Health. 2024;12 (3):e382–e395.

16 Tromp J, Ferreira JP, Janwanishstaporn S, et al. Heart failure around the world. Eur J Heart Fail. 2019;21(10):1187–1196.

17 Gupta A, Tisdale RL, Calma J, et al. Equity in the Setting of Heart Failure Diagnosis: An Analysis of Differences Between and Within Clinician Practices. Circ Heart Fail. 2024; 17(6):e010718.

18 GBD 2019 Demographics Collaborators. Global age-sex-specific fertility, mortality, healthy life expectancy (HALE), and population estimates in 204 countries and territories, 1950 -2019: a comprehensive demographic analysis for the Global Burden of Disease Study 2019. Lancet. 2020;396(10258):1160–1203.

19 Mitsnefes MM. Cardiovascular disease in children with chronic kidney disease. J Am Soc Nephrol. 2012;23(4):578–585.

20 Regitz-Zagrosek V. Sex and sex Differences in Heart Failure. Int J Heart Fail. 2020;2 (3):157–181.

21 Khan SS, Beach LB, Yancy CW. Sex-Based Differences in Heart Failure: JACC Focus Seminar 7/7. J Am Coll Cardiol. 2022;79(15):1530–1541.

22 Conrad N, Judge A, Tran J, et al. Temporal trends and patterns in heart failure incidence: a population-based study of 4 million individuals. Lancet. 2018;391(10120):572–580.

23. GBD Chronic Kidney Disease Collaboration. Global, regional, and national burden of chronic kidney disease, 1990-2017: a systematic analysis for the Global Burden of Disease Study 2017. Lancet. 2020;395(10225):709–733.

24 Khatib R, McKee M, Shannon H, et al. Availability and affordability of cardiovascular disease medicines and their effect on use in high-income, middle-income, and low-income countries: an analysis of the PURE study data. Lancet. 2016;387(10013):61–69.

25 Echouffo-Tcheugui JB, Zhang S, Florido R, et al. Duration of Diabetes and Incident Heart Failure: The ARIC (Atherosclerosis Risk In Communities) Study. JACC Heart Fail. 2021; 9(8):594–603.

26 Darenskaya M, Kolesnikov S, Semenova N, Kolesnikova L. Diabetic Nephropathy: Significance of Determining Oxidative Stress and Opportunities for Antioxidant Therapies. Int J Mol Sci. 2023;24(15):12378. Published 2023 Aug 3.

27 Burnier M, Damianaki A. Hypertension as Cardiovascular Risk Factor in Chronic Kidney Disease. Circ Res. 2023;132(8):1050–1063.

28 Messerli FH, Rimoldi SF, Bangalore S. The Transition From Hypertension to Heart Failure: Contemporary Update [published correction appears in JACC Heart Fail. 2017 Dec;5(1 2):948.

